# Trans auricular vagus nerve stimulation for aneurysmal sub arachnoid haemorrhage (VNS-SAH): a pilot randomised controlled trial

**DOI:** 10.64898/2026.08.25.26361366

**Authors:** Matthew Myers, Finlay Robson, Sheharyar Baig, Saminderjit Kular, Mudasar Aziz, Elisabetta Burchi, Debapriya Bhattacharyya, Li Su, Arshad Majid, Ali N Ali

**Author notes:** **Corresponding authors:** Prof Arshad Majid Department of Neurosciences, School of Medicine and Population Health, University of Sheffield, UK., Dr Ali N Ali, Department of Neurosciences, School of Medicine and Population Health, University of Sheffield, UK.

## Abstract

**Background:** Aneurysmal subarachnoid haemorrhage (aSAH) is frequently complicated by delayed cerebral ischaemia (DCI), for which current therapies incompletely target the underlying multifactorial pathophysiology. Transauricular vagus nerve stimulation (taVNS) modulates inflammatory, vasoactive and autonomic pathways and may attenuate secondary brain injury after aSAH.

**Methods:** We conducted a prospective, single-centre, single-blind, randomised, sham-controlled pilot trial in adults within 5 days of aneurysm securing for non-traumatic aSAH. Participants were allocated 1:1 to active taVNS (left tragus) or sham (left earlobe) using a portable device delivered for 45 minutes twice daily over 5 days. Primary outcomes were safety (taVNS-related serious adverse events), acceptability, and compliance; secondary outcomes included inflammatory biomarkers, DCI, in-hospital complications, and functional outcomes to 1 month.

**Results:** Thirty patients were randomised (16 taVNS, 14 sham), with numerically more severe aSAH at baseline in the taVNS arm. No taVNS-related serious adverse events occurred; side effects were generally mild and transient, and over 80% of planned sessions were completed. TaVNS produced greater reductions in serum tumour necrosis factor-α and trends towards reductions in interleukin-1β and interleukin-10, with numerically fewer DCI events (6.6% vs 35.7%) and neurological impairments (16.7% vs 53.8%), although functional outcomes were not statistically different at 1 month.

**Conclusions:** Early taVNS after aSAH is safe, acceptable, and feasible in the neurocritical care setting and shows biologically plausible signals warranting evaluation in larger multicentre trials.

## Introduction

Aneurysmal subarachnoid haemorrhage (aSAH) occurs following rupture of an intracranial aneurysm, and accounts for approximately 7% of all strokes [1]. Estimates indicated 30-day mortality rates of approximately 20% [2], with nearly half of individuals left with permanent disability, despite timely aneurysm securing [1]. In addition to the immediate impact of bleeding into the subarachnoid space, poor outcomes are heavily influenced by secondary brain injury that develops in the days and weeks following ictus including large and small vessel vasospasm, delayed (DCI), hydrocephalus and seizures [3]. DCI in particular is a sequalae of aSAH which occurs in up to 30% of patients usually within 14 days of ictus, and alongside rebleeding is the strongest predictor of in hospital mortality [4].

The pathophysiology of DCI is increasingly recognised as multifactorial, encompassing large vessel vasospasm, microvascular dysfunction, localised coagulopathy, cortical spreading ischaemia, breakdown of the blood brain barrier, local and systemic inflammatory responses and an impairment of cerebral autoregulation [5]. Current management strategies focus predominantly on haemodynamic augmentation and nimodipine to counteract vasospasm [6], however this fails to address other important factors in the disease pathophysiology. Interventions that target multiple pathophysiological pathways may offer greater clinical benefits.

Vagus nerve stimulation (VNS), and its non-invasive counterpart transauricular vagus nerve stimulation (taVNS) are non-pharmacological strategies that may modulate these diverse processes. In pre-clinical studies, delivery of taVNS in the acute period following experimental stroke has been shown to reduce systemic inflammatory cytokine surges through effects on alpha 7 nicotinic acetylcholine receptor interactions [7], that also facilitates polarisation of microglia centrally from a M1 (proinflammatory) to a M2 (anti-inflammatory) state [8]. Further, short bursts of VNS can reduce cortical spreading depolarisations (a feature of excitotoxicity) [9], reduce expression of matrix metalloproteinases involved in BBB breakdown [10] and lead to vasoactive and angiogenic effects that promote cerebral perfusion [11].

TaVNS represents a practical neuromodulatory intervention that can be delivered at the bedside and initiated soon after aneurysm securing, simultaneously targeting inflammatory, autonomic and cerebral hemodynamic pathways in DCI. This pilot study aimed to assess the safety, tolerability and feasibility of taVNS as a treatment for aSAH and explored its effect on rates of DCI and measures of functional recovery.

## Methods

This study was a prospective, single centre, single blind randomised sham-controlled trial with blinded outcome assessment. The study was designed with the Sheffield Stroke and Aphasia Interest Group and approved by the Health Research Authority and Health and Care Research Wales Ethics Committee (24/YH/0024) and registered with ClinicalTrials.gov (NCT06374693). Adult patients admitted to Sheffield Teaching Hospitals with confirmed non-traumatic aSAH were screened for enrolment. Inclusion criteria included: 1) age ≥ 18 years; 2) angiographic confirmation of aSAH; 3) within 5 days of emergency aneurysm securing procedure (coiling or clipping). Current or prior use of a vagus nerve stimulation device, history of second- or third-degree heart block or ECG confirmed bradycardia (heart rate < 40 per minute), presence of other implanted electrical stimulator, and pregnancy where exclusions to participation. Written consent was taken from participants who had the capacity to make decisions about their participation, for those who lacked capacity, assent was obtained from the participants legal representatives. A study flow diagram is depicted in Figure 1. Participants were block randomised (1:1) using an online system (Sealed Envelope Ltd 2017), stratified according to age (<65 years and ≥ 65 years) to receive either active taVNS or sham by an independent researcher. The participants, their families, the treating medical teams and the research outcome assessors were blinded to treatment allocation, however research staff administering the intervention were unblinded.

Intervention - Active taVNS was delivered using the Neurosym^®^ device, applied to the tragus of the left ear for 45 minutes, twice daily (AM and PM), for 5 days (10 sessions total). Stimulation parameters included pulse width 25ms, frequency 20 Hz, and intensity was set to below pain threshold in alert participants, and 25mA in those who were intubated and ventilated. Sham taVNS was exactly the same except stimulated the earlobe to avoid the auricular branch of the vagus nerve.

Outcome measures were collected at baseline, end of treatment (EOT), discharge and 1 month by researchers blinded to treatment allocation and intervention delivery. Baseline socio-demographic and clinical characteristics were recorded, including Hunt and Hess scale (H&H) of aSAH severity [13]. Primary outcomes of safety, acceptability and compliance were recorded during the intervention and follow up period by the participants responsible medical teams. To assess safety, adverse events or serious adverse events either related or unrelated to the intervention were systematically and prospectively recorded. Safety was predefined as no SAEs related to taVNS. Acceptability was measured by asking participants to rate their experience off several expected side effects (pain or paraesthesia, headache, nausea or vomiting, coryza, dizziness, facial drooping) during the intervention sessions on a 5-point Likert scale (1=none, 5=extremely severe) after each treatment session. Acceptability was defined as less than 1/3 of participants reporting moderate or greater discomfort (mean score ≥ 3). Treatment logs were used to measure compliance and predefined as completion of more than 80% of intended sessions. Secondary outcome measures included inpatient mortality (%), length of hospital stay (days), and rates (%) of new onset (post randomisation) seizures, rebleeding, hydrocephalus, hyponatremia (< 140 mmol/L) and DCI as defined by the subarachnoid haemorrhage international trialists (new focal neurological impairment or decrease in Glasgow Coma Score of ≥ 2 points, lasting > 1 hour and not explained by any other cause e.g. hydrocephalus, infection, or new confirmed cerebral infarction) [12]. A radiologist blinded to treatment allocation reviewed all neuroimaging. Modified Ranking score (mRS), Barthell index (BI) and headache scores (Likert 1-10; 1=none, 10=extremely severe) were recorded at discharge and 1 month. Blood samples were taken at baseline and EOT, and serum processed immediately and stored for analysis of key inflammatory mediators IL-1β, IL-6, IL-10 and TNF-α.

Data were analysed using IBM SPSS statistics V 29. We aimed to recruit a minimum of 12 participants per group required to assess feasibility of an intervention and provide data to estimate sample sizes for future powered definitive studies [14]. Baseline demographic and clinical characteristics were reported descriptively as were data on safety, acceptability and compliance. Exploratory, intention-to-treat analyses to assess between group differences in baseline characteristics and for secondary outcomes employed Mann-Whitney U or Chi square tests (non-normally distributed data) where appropriate (2-sided p taken at < 0.05), and 1-month mRS ordinal shift analysis, adjusted for baseline severity, was undertaken [15].

## Results

From April 2024 to September 2025, 73 patients were approached and 30 participants recruited (taVNS 16: sham 14) into the study. Only 1 participant in the taVNS group withdrew prior to end of treatment, meaning that 1 month follow up data was available for 29 individuals (96.7%). Participant characteristics are detailed in Table 1. Nearly three quarters of participants were female. The taVNS group consisted of numerically but not statistically higher grades of aSAH (H&H, increased number intubated and ventilated) than the sham group. All participants were treated with nimodipine. Paired baseline and EOT inflammatory marker analyses were available for 23 participants (taVNS = 11, sham = 12).

**Table 1.**
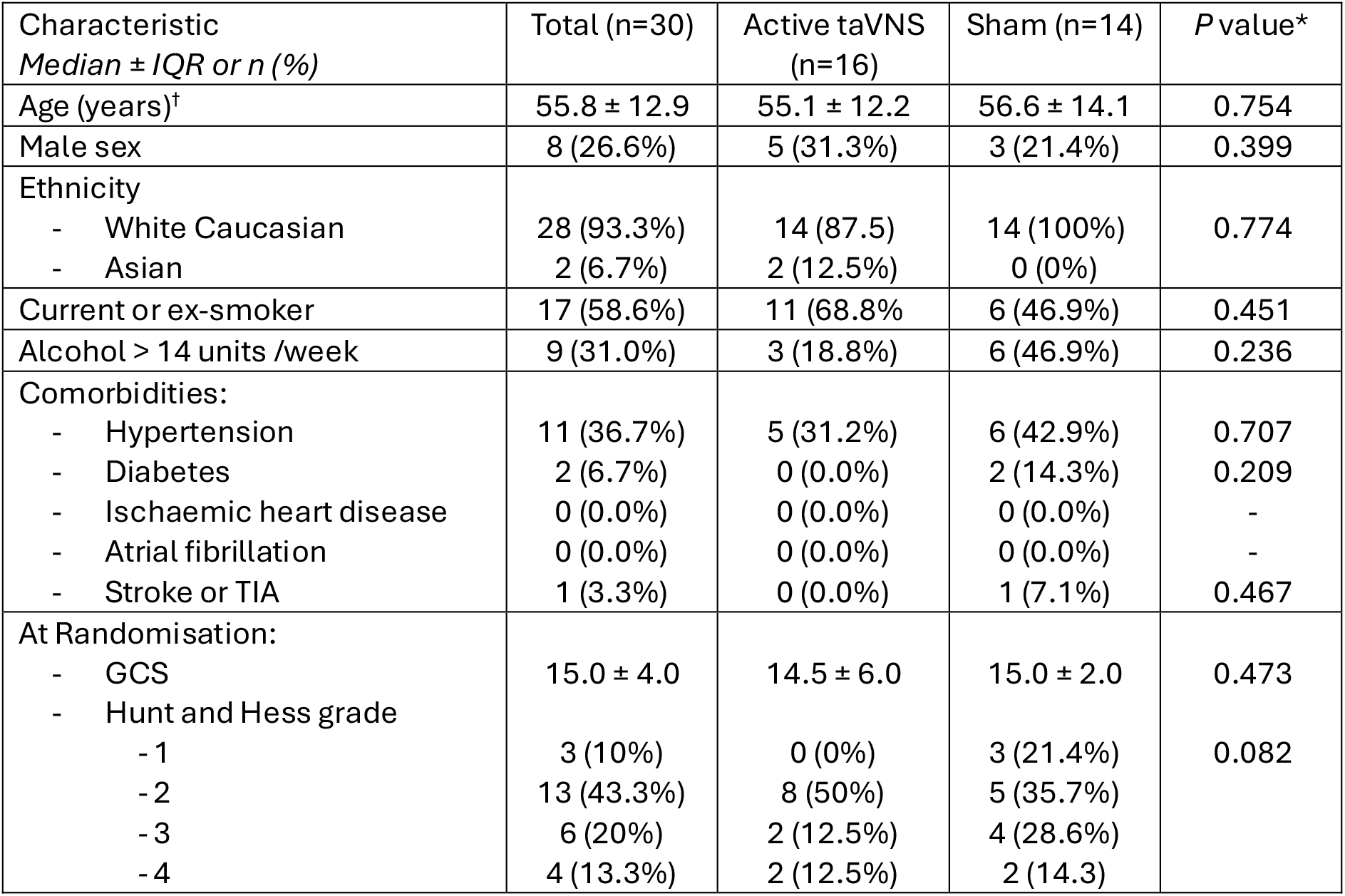
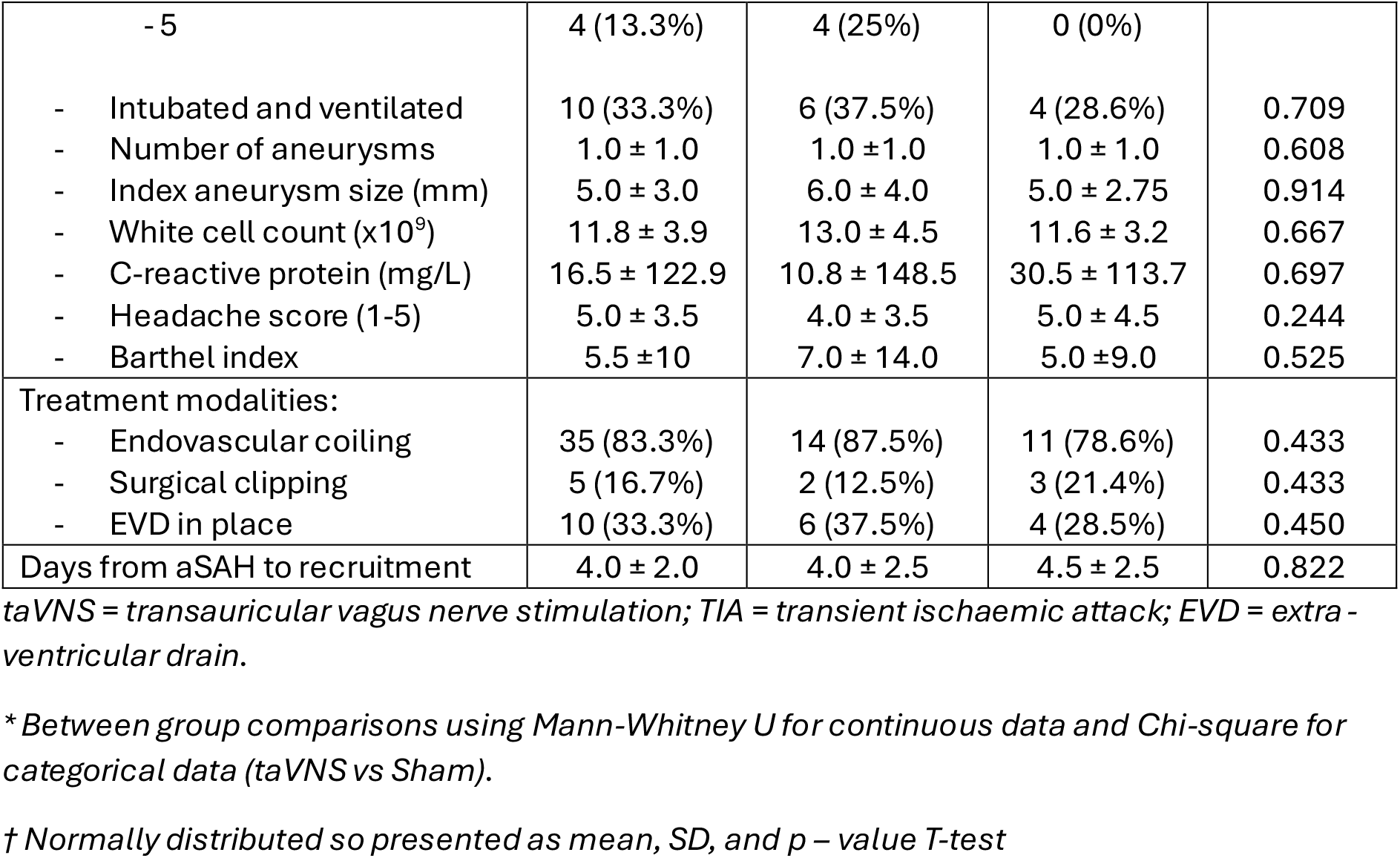
Baseline characteristics of study participants.

| Characteristic<br>*Median ± IQR or n (%)* | Total (n=30) | Active taVNS<br>(n=16) | Sham (n=14) | *P* value* |
| --- | --- | --- | --- | --- |
| Age (years)<sup>†</sup> | 55.8 ± 12.9 | 55.1 ± 12.2 | 56.6 ± 14.1 | 0.754 |
| Male sex | 8 (26.6%) | 5 (31.3%) | 3 (21.4%) | 0.399 |
| Ethnicity |  |  |  |  |
| - White Caucasian | 28 (93.3%) | 14 (87.5) | 14 (100%) | 0.774 |
| - Asian | 2 (6.7%) | 2 (12.5%) | 0 (0%) |  |
| Current or ex-smoker | 17 (58.6%) | 11 (68.8%) | 6 (46.9%) | 0.451 |
| Alcohol > 14 units /week | 9 (31.0%) | 3 (18.8%) | 6 (46.9%) | 0.236 |
| Comorbidities: |  |  |  |  |
| - Hypertension | 11 (36.7%) | 5 (31.2%) | 6 (42.9%) | 0.707 |
| - Diabetes | 2 (6.7%) | 0 (0.0%) | 2 (14.3%) | 0.209 |
| - Ischaemic heart disease | 0 (0.0%) | 0 (0.0%) | 0 (0.0%) | - |
| - Atrial fibrillation | 0 (0.0%) | 0 (0.0%) | 0 (0.0%) | - |
| - Stroke or TIA | 1 (3.3%) | 0 (0.0%) | 1 (7.1%) | 0.467 |
| At Randomisation: |  |  |  |  |
| - GCS | 15.0 ± 4.0 | 14.5 ± 6.0 | 15.0 ± 2.0 | 0.473 |
| - Hunt and Hess grade |  |  |  | 0.082 |
| - 1 | 3 (10%) | 0 (0%) | 3 (21.4%) |  |
| - 2 | 13 (43.3%) | 8 (50%) | 5 (35.7%) |  |
| - 3 | 6 (20%) | 2 (12.5%) | 4 (28.6%) |  |
| - 4 | 4 (13.3%) | 2 (12.5%) | 2 (14.3%) |  |
| - 5 | 4 (13.3%) | 4 (25%) | 0 (0%) |  |
| - Intubated and ventilated | 10 (33.3%) | 6 (37.5%) | 4 (28.6%) | 0.709 |
| - Number of aneurysms | 1.0 ± 1.0 | 1.0 ± 1.0 | 1.0 ± 1.0 | 0.608 |
| - Index aneurysm size (mm) | 5.0 ± 3.0 | 6.0 ± 4.0 | 5.0 ± 2.75 | 0.914 |
| - White cell count (x10<sup>9</sup>) | 11.8 ± 3.9 | 13.0 ± 4.5 | 11.6 ± 3.2 | 0.667 |
| - C-reactive protein (mg/L) | 16.5 ± 122.9 | 10.8 ± 148.5 | 30.5 ± 113.7 | 0.697 |
| - Headache score (1-5) | 5.0 ± 3.5 | 4.0 ± 3.5 | 5.0 ± 4.5 | 0.244 |
| - Barthel index | 5.5 ± 10 | 7.0 ± 14.0 | 5.0 ± 9.0 | 0.525 |
| Treatment modalities: |  |  |  |  |
| - Endovascular coiling | 35 (83.3%) | 14 (87.5%) | 11 (78.6%) | 0.433 |
| - Surgical clipping | 5 (16.7%) | 2 (12.5%) | 3 (21.4%) | 0.433 |
| - EVD in place | 10 (33.3%) | 6 (37.5%) | 4 (28.5%) | 0.450 |
| Days from aSAH to recruitment | 4.0 ± 2.0 | 4.0 ± 2.5 | 4.5 ± 2.5 | 0.822 |

taVNS = transauricular vagus nerve stimulation; TIA = transient ischaemic attack; EVD = extra-ventricular drain.
\* Between group comparisons using Mann-Whitney U for continuous data and Chi-square for categorical data (taVNS vs Sham).
† Normally distributed so presented as mean, SD, and p – value T-test

Safety, acceptability and compliance - No participants in either group experienced any SAE related to the intervention. A total of 4/30 participants died during their inpatient stay (13.3%), 3 in the taVNS group (mean H&H score 4.7) and 1 in the sham group (H&H score 3). One participant in the taVNS group withdrew after three treatments due to headache, although their pre-treatment headache score was high (Likert 7/10). During the intervention period headache was reported by 65% of participants alert enough to provide feedback (40% taVNS vs 90% sham), 30% reported paraesthesia (40% taVNS vs 20% sham) and 20% nausea (0% taVNS vs 40% sham). Expected device side effects were mild and transitory. Ony 30% of participants reported side effect severity scores of ≥ 3 at any point. Taking into account death before the end of the treatment and early discharge from hospital, a total of 259 possible treatment sessions were possible. In both groups, a total of 221 (85.3%) sessions were delivered, 215 of which (83%) were the full 45 minutes. Mean (SD) stimulation intensity in the taVNS group 24.4 (4.8) was similar to the sham group 23.9 (4.3).

Between group differences in secondary outcomes are highlighted in Table 2. At EOT taVNS was associated with significant reductions in serum TNF-α, and trends towards reductions in IL-1β, and IL-10 (Figure 2). No statistically significant differences in any other secondary outcomes were observed, either at discharge or 1 month follow up, however, numerically fewer participants in the taVNS group experienced DCI and were left with ongoing neurological impairments. When evaluating ordinal shift of mRS, an ordinal logistic model adjusting for H&H grade at baseline showed the common odds ratio for better mRS at 1 month favoured taVNS but was not statistically significant (OR 0.83, 95%CI 0.16 to 4.24; p-0.82).

**Table 2.** Secondary outcomes measures at end of treatment, discharge and 1 month follow up.

| Outcome<br>Median ± IQR or n (%) | taVNS (n=15) | Sham (n=14) | P-value |
| --- | --- | --- | --- |
| **End of treatment:** |  |  |  |
| Change in serum inflammatory mediators* (baseline to end of treatment): |  |  |  |
| - IL-1β (pg/ml) | -371 ± 572 | +684 ± 1562 | 0.05 |
| - IL-6 (pg/ml) | -1406 ± 17496 | -3679 ± 7497 | 0.798 |
| - IL-10 (pg/ml) | -349 ± 1721 | +809 ± 2372 | 0.05 |
| - TNF-α (pg/ml) | -373 ± 1151 | +504 ± 1535 | 0.028 |
| - CRP (mg/L) | -29.1 ± 139.2 | -24.2 ± 114.2 | 0.762 |
| Change in WCC (x10<sup>9</sup>) | -3.2 ± 3.6 | -3.3 ± 2.4 | 0.961 |
| **At discharge:** |  |  |  |
| Inpatient mortality | 3 (20%) | 1 (7%) | 0.351 |
| Length of hospital stay (days)** | 9.0 ± 14.5 | 18.0 ± 12.0 | 0.193 |
| Neurological complications: |  |  |  |
| - New seizures | 0 (0%) | 0 (0%) | NA |
| - Rebleeding | 0 (0%) | 0 (0%) | NA |
| - Hydrocephalus | 1 (6.6%) | 0 (0%) | 0.533 |
| - Hyponatraemia | 2 (13.3%) | 4 (28.6%) | 0.261 |
| - DCI | 1 (6.6%) | 5 (35.7%) | 0.059 |
| Ongoing focal neurological impairment<sup>†</sup> | 2 (16.7%) | 7 (53.8%) | 0.101 |
| Headache score (1-10) | 6.0 ± 8 | 2.0 ± 5 | 0.755 |
| Barthel index | 20.0 ± 4.5 | 17.0 ± 4.5 | 0.244 |
| mRS | 2.0 ± 3.75 | 2.0 ± 1.0 | 0.854 |
| **At 1 month follow up:** |  |  |  |
| Mortality | 3 (20%) | 2 (14%) | 0.567 |
| Headache score (1-10) | 4.5 ± 6 | 2.0 ± 6 | 0.449 |
| Barthel index | 20.0 ± 3.0 | 20.0 ± 0.0 | 0.424 |
| mRS | 2.0 ± 1.0 | 1.0 ± 1.0 | 0.244 |
| - Good outcome (mRS 0-2) | 75% | 77% |  |
| - Poor outcome (mRS 3-6) | 25% | 23% |  |

\* N=23 samples (12 taVNS vs 11 sham)
\*\*Excluding those dying as inpatients
† Of those still alive at discharge.
DCI = delayed cerebral ischaemia; IL-1β = Interleukin 1 $\beta$ ; IL-6 = Interleukin 6; IL-10 = Interleukin 10; TNF-α = Tumour necrosis factor alpha

## Discussion

This pilot randomised controlled trial demonstrates that taVNS initiated within 5 days of aneurysm securing in aSAH is safe, acceptable and feasible with high protocol adherence and no taVNS related SAEs despite treatment in a high acuity neurocritical care environment. Side effect profiles were mild and transient and overall compliance exceeded 80% of planned sessions. These data align with the safety and acceptability profiles seen in two recently published randomised controlled trials [16,17], as well as acute ischaemic and haemorrhagic stroke cohorts [18,19].

Although not powered for efficacy, taVNS was associated with biologically plausible modulation of systemic inflammation. Compared with sham, taVNS produced greater reductions in TNF-α, and a trend towards reductions in IL-1β and IL-10. Elevated concentrations of TNF-α, IL-6 and indeed CRP after aSAH have been linked to worse long-term functional outcomes in observational cohorts, even after adjustment for initial clinical severity [20]. Huguenard et al similarly randomised 27 individuals with aSAH to 7 days (20 minutes twice daily) of either taVNS delivered at the concha (n=13) or earlobe stimulation sham (n=14). They also demonstrated significant reductions in serum TNF-α, but also IL-6 from treatment initiation up to 14 days [16]. We also found taVNS was associated with a trend towards a reduction in circulating levels of the potent anti-inflammatory IL-10, which usually acts to suppress cytokines such as TNF-α and IL-1β [21], thus the interplay between taVNS and inflammatory mediators requires further exploration. Whether related to modulation of systemic inflammation, or direct vasoactive effects [11], we observed numerically fewer cases of DCI and subsequent neurological impairment in the taVNS arm. These exploratory signals are consistent with the findings of Huguenard et al who demonstrated significant reductions in rates of radiologically confirmed vasospasm and poor clinical outcomes in patients treated with taVNS compared to sham [16]. Further, Rebeiz et al randomised 40 patients with all types of SAH to either transcervical VNS (tcVNS) delivered for 2 mins 4 times daily (n=19), or sham (n=21) for a median of 9 days, and found numerically fewer rates of new cerebral infarction in the tcVNS group (5.6% vs 23.8%) [17]. Taken together with preclinical and early clinical stroke data, these trials support a unifying model whereby taVNS modulates the cholinergic anti-inflammatory pathway and central autonomic networks that reduce secondary brain injury after aSAH.

Several limitations temper interpretation of this data. This was a single centre, small pilot study. The TAVNS group had numerically higher baseline H&H grades and inflammatory markers, which may have expected to bias against detecting benefit. Cytokine measurements were limited to two time points and are susceptible to confounding by intercurrent infection, surgical complications and hydrocephalus [20]. Our sample size and short (1 month) follow up precludes firm conclusions about DCI, neurological disability and durable functional outcomes, and we did not incorporate radiological imaging to confirm changes to vessel calibre that may have evaluated vascular mechanisms of action. However loss to follow up was low, and outcome assessment blinded, enhancing confidence in the data collected.

## Conclusion

TaVNS initiated early after aneurysm securing in aSAH was safe, well tolerated and deliverable at scale in the neurocritical care setting, and demonstrated beneficial signals of effect on systemic inflammation, rates of DCI and neurological impairment. Future multicentre, adequately powered, RCTs with longer term functional follow up are warranted to determine if taVNS can improve clinically meaningful outcomes after aSAH.

## Data Availability

Data are available upon reasonable request.

